# Prediction of Multiple Individual Primary Cardiovascular Events Using Pooled Cohorts

**DOI:** 10.1101/2023.08.01.23293525

**Authors:** Jeremy B. Sussman, Rachael T. Whitney, James F. Burke, Rodney A. Hayward, Andrzej Galecki, Stephen Sidney, Norrina Bai Allen, Rebecca F. Gottesman, Susan R. Heckbert, William T. Longstreth, Bruce M Psaty, Mitchell S.V. Elkind, Deborah A. Levine

## Abstract

**Introduction:** Most current clinical risk prediction scores for cardiovascular disease prevention use a composite outcome. Risk prediction scores for specific cardiovascular events could identify people who are at higher risk for some events than others informing personalized care and trial recruitment. We sought to predict risk for multiple different events, describe how those risks differ, and examine if these differences could improve treatment priorities.

**Methods:** We used participant-level data from five cohort studies. We included participants between 40 and 79 years old who had no history of myocardial infarction (MI), stroke, or heart failure (HF). We made separate models to predict 10-year rates of first atherosclerotic cardiovascular disease (ASCVD), first fatal or nonfatal MI, first fatal or nonfatal stroke, new-onset HF, fatal ASCVD, fatal MI, fatal stroke, and all-cause mortality using established ASCVD risk factors. To limit overfitting, we used elastic net regularization with alpha = 0.75. We assessed the models for calibration, discrimination, and for correlations between predicted risks for different events. We also estimated the potential impact of varying treatment based on patients who are high risk for some ASCVD events, but not others.

**Results:** Our study included 24,505 people; 55.6% were women, and 20.7% were non-Hispanic Black. Our models had C-statistics between 0.75 for MI and 0.85 for HF, good calibration, and minimal overfitting. The models were least similar for fatal stroke and all MI (0.58). In 1,840 participants whose risk of MI but not stroke or all-cause mortality was in the top quartile, we estimate one blood pressure-lowering medication would have a 2.4% chance of preventing any ASCVD event per 10 years. A moderate-strength statin would have a 2.1% chance. In 1,039 participants who had top quartile risk of stroke but not MI or mortality, a blood pressure-lowering medication would have a 2.5% chance of preventing an event, but a moderate-strength statin, 1.6%.

**Conclusion:** We developed risk scores for eight key clinical events and found that cardiovascular risk varies somewhat for different clinical events. Future work could determine if tailoring decisions by risk of separate events can improve care.

## Background

Risk prediction is a the key element of all treatment recommendations in cardiovascular primary prevention.^1–5^ In particular, the risk score developed from the Pooled Cohort Equations (PCEs) is at the center of primary prevention recommendations for cholesterol reduction, blood pressure (BP) treatment, and aspirin use.^2, 6^ The PCEs were a substantial advance. By combining multiple populations, all with well-validated data, they were based on a wealth of evidence that previous cardiovascular risk scores lacked.

One key limitation of the PCEs is that they only predict a single composite outcome – primary major atherosclerotic cardiovascular disease (ASCVD) events, defined as myocardial infarction (MI) or stroke among participants who have never had one before. Developing scores to predict distinct multiple separate cardiovascular outcomes, such as MI, stroke, heart failure (HF), ASCVD mortality, and total mortality, could be useful, especially since treatments are not uniformly effective across these events. For example, since BP reduction has a larger effect on stroke than on MI and low-density lipoprotein (LDL) cholesterol lowering has a larger effect on MI than stroke,^7, 8^ identifying patients at especially high risk for specific event types might improve treatment decisions. In fact, accounting for these types of differences could improve health through multiple mechanisms, most obviously, by enabling more effective tailoring of treatment approaches to individual people. Predicting specific outcome types could also help since many people struggle to understand composite risk scores and may have greater fear for some event types, such as a strong desire to never have a stroke. Finally, these scores could also be used in decision analysis and cost-effectiveness analyses whenever a new treatment is more effective at preventing one type of ASCVD event than another.

In this study we used individual participant data from five well-characterized US cardiovascular cohorts to create risk scores for multiple clinical event types. We also assessed the risk scores for reliability and accuracy, examined correlations between risk for the different event types, and examined the differences in participants at risk for the different event types. Specifically, we sought to assess if we could predict first ASCVD, first MI, first stroke, new onset HF, fatal ASCVD, fatal MI, fatal stroke, and all-cause mortality; if we could identify participants who have meaningfully different risk for one type of event than others; and if we could estimate how these distinctions might alter treatment priorities.

## Methods

### General

This is a study from the larger BP COG study, which was designed to understand the relationships between BP levels and cognitive decline in Black, White, and Hispanic participants. BP COG analyzed pooled individual participant data from large, high-quality NIH-funded cohort studies, some of which overlap with those in the PCEs, specifically Atherosclerosis Risk in Communities Study (ARIC), Cardiovascular Health Study (CHS), and Framingham Offspring Study (FOS). Our dataset also includes the Northern Manhattan Study (NOMAS) and Multi-Ethnic Study of Atherosclerosis (MESA), which provide two populations of Latino participants. It does not include the original Framingham study, which was conducted in a period when ASCVD rates and case ascertainment were different from today, or Coronary Artery Risk Development in Young Adults, for which the baseline participant age was below our studies’ age eligibility. Data were collected from January 1971 to December 2019.

### Population

Participants ≥ 40 and <80 years of age at baseline, who were Black, White, or Hispanic, and had no history of MI, stroke, or HF were included in this study. Our process of harmonizing cohort data has been previously described.^9–11^

### Outcomes

Our outcome variables were first ASCVD, first MI (including fatal or nonfatal), first stroke (including fatal or nonfatal), new onset HF, fatal MI, fatal stroke, fatal ASCVD and all-cause mortality occurring within 10 years of baseline assessment. These event types are biologically related, of high public health importance, and often combined into single composite scores. All-cause mortality was included because of its general importance. Since first ASCVD, first MI, and first stroke all included both fatal and nonfatal events, there was substantial overlap between many of the outcome variables. We used 10 years of follow-up because that is what was used in the PCEs, the most important risk score in current ASCVD clinical practice.^2, 6^

### Predictors

Predictor variables associated with the events under study in previous research were chosen.^6, 12, 13^ They included age (years), gender (female vs male), race or ethnicity (non-Hispanic Black, Hispanic, or non-Hispanic White), tobacco use (current, former, or never), body-mass index (kg/m²), low-density lipoprotein (LDL) cholesterol (mg/dL), on cholesterol medications, history of diabetes, systolic BP (mmHg), on BP medications, history of atrial fibrillation (no vs yes) and estimated glomerular filtration rate (mL/min/1.73m²). We also looked at interaction terms for gender by age, gender by systolic BP, race/ethnicity by age, systolic BP by on BP medications, LDL cholesterol by on cholesterol medications, and age-squared. Interaction terms were selected based on previous research.^6, 12, 13^ Race and ethnicity are included as imperfect markers of complex economic and sociocultural phenomena, not as biological variables. Gender is included as a combination of both biological and social variables.

### Analysis

For our primary analyses we used logistic regression with elastic net regularization (ENR) with alpha set at 0.75. ENR is like traditional regression models but with added elements to reduce overfitting, which is where the model attributes to prediction what is actually due to chance. In ENR this reduction is accomplished by shrinking the observed predictions, either by assuming the true predictive effect of a variable is smaller than that which is observed or by removing from the predictive model variables that might improve prediction by a small amount, on the likelihood that the benefit is only due to chance. The elastic net model does not show p-values for individual predictor variables, but variables with effects smaller than a threshold based on the selected alpha are removed from the model. The alpha is a way of choosing which shrinkage technique to prioritize, with 0.75 reflecting our team’s decision to slightly prioritize removing variables from the model to yield a smaller, more parsimonious model.

Unlike many existing models, we did not separate our models by race/ethnicity and gender. We did this to minimize overfitting, because of existing research that this approach is more effective and because we do not believe the biology of race, ethnicity, or gender merits that separation.^14, 15^

To see how similar risk is between cardiovascular conditions, we compared predicted risks from across all models using correlations. Then, to examine the clinical differences between participants for whom our models gave a high predicted risk for different conditions and the impact of treatment on their observed events, we identified participants in the top quartile of risk for multiple measures and examined how they differed from those in the top quartile for other event types. We used this to estimate the likely clinical benefit of ASCVD reduction from one moderate dose BP medicine vs. one moderate potency statin medicine for these groups of “isolated high risk.” BP medications reduce stroke rates more than MI rates and statin medicines lower the rates of each similarly. ^7, 8^ Therefore, people who are high stroke risk will have greater benefit from BP medications than would be expected by ASCVD risk alone. We hypothesized that we could identify people who are high stroke risk and that this would differentially guide medication management to prevent more events with less medication use. We used estimates of relative risk reduction from the Trialists Treatment Collaboratives, estimating that a single moderate dose BP medicine lowers systolic BP by 6.3 mmHg and a 5% reduction in BP lowers stroke rate by 19% and MI rate by 6.3%. We estimated that a single moderate potency statin medicine lowers stroke rate by 13% and MI rate by 14%.^7, 8^

One potentially important analytic concern was between-cohort heterogeneity. Different cohorts can identify different event rates for participants with the same characteristics. This limited external validity is most likely caused by an ascertainment bias, in which some cohorts identified events that others may have missed, such as minor MIs. To address this possibility, we developed a technique to normalize the results between each cohort. First, we ran the models for each CVD outcome with a variable that identifies the cohort. Next, we converted the beta-parameters for each cohort variable to zero, so that differences that are attributable purely to cohort phenomenon were removed. We then set a Y-intercept (beta-zero) to a value that yielded the same number of predicted events as in the original model. The effect of this approach is to predict the same number of events but remove the variability due to specific cohort effects.

### Evaluation

All predictive models were first evaluated with visual inspection of the predicted plots.^16^ We tested discrimination using the C-statistic (measuring the likelihood that higher risk participants are more likely to have the outcome); calibration using calibration slope, graphically (measuring how the predicted event rates match observed rates without consistent over- or under-prediction); the ability to stratify individual participant risk using interquartile range; and the calibration-in-the-large (comparing the overall mean event rate with the overall predicted mean event rate). We assessed overall accuracy using the Brier score (measuring overall accuracy of prediction, in which larger errors are weighted more strongly) and internal validity. Our primary internal validity check was a form of cross-validation in which we derived models on 80% of the sample and then tested them on both the derivation sample and the remaining 20% validation sample. We did this 10 times for each model to understand the variability of the results. The validation results are the primary results. The derivation samples were retained to observe how different it was from the validation results. A larger change is a marker of overfitting.

### Sensitivity analyses

We performed two sensitivity analyses. The first used logistic regression without variable selection instead of regression penalized with ENR. In theory ENR will minimize overfitting, which will make it more effective when the sample size is small relative to the number of predictor variables. ENR is, however, less easily available and takes much longer computational time. The second sensitivity analysis looked at using 5-year risk scores instead of 10. Ten-year scores are more common in ASCVD research and practice, but having enough follow-up is not always practical and it’s possible that 10 years of follow-up doesn’t reflect that person’s immediate needs as well as 5 years.

## Results

Our sample’s mean age was 58.6 and was 55.6% women. The mean BP was 138.4. Of all participants, 20.7% were non-Hispanic Black and 10.2% were Hispanic (Table 1). Almost half the people in the study (47.4%) were from the ARIC cohort.

**Table 1:** Baseline characteristics.

| **Table 1: Baseline characteristics** |  |
| --- | --- |
| **Characteristics** | **Participants**<br>**(n=24,505)** |
| Age, mean (SD), y | 58.6 (9.7) |
| Women, n,% | 13,627 (55.6) |
| Tobacco use, n,% | 5,308 (21.7) |
| Systolic Blood Pressure, mean<br>(SD), mmHg | 138.4 (20.6) |
| BMI, mean (SD), kg/m<sup>2</sup> | 27.7 (5.2) |
| LDL cholesterol, mean, (SD),<br>mg/dL | 131.5 (37.4) |
| Diabetes, n,% | 2,415 (10.0) |
| eGFR, mean (SD), mL/min | 74.9 (17.2) |
| Race/Ethnicity |  |
| Non-Hispanic Black, n,% | 5,084 (20.7) |
| Non-Hispanic White, n,% | 16,924 (69.1) |
| Hispanic, n,% | 2,497 (10.2) |
| Study |  |
| ARIC, n, % | 11,625 (47.4) |
| CHS, n, % | 3,755 (15.3) |
| FOS, n, % | 1,801 (7.4) |
| MESA, n, % | 5,613 (23.0) |
| NOMAS, n, % | 1,709 (7.0) |

Our primary models for all 10-year risk predictions are presented in Table 2. White race was not independently associated with MI, HF, or fatal MI, but was negatively associated with stroke, ASCVD, all-cause mortality, fatal ASCVD, and fatal stroke, compared to non-Black Hispanic ethnicity. Black race was associated with increased stroke and MI rates. Both higher systolic BP and being on BP lowering medicine were associated with increased risk of every outcome. We did identify interaction effects between race/ethnicity and other risk factors. In particular, the impact of BP was greater in Black participants for almost all outcomes, as indicated by positive Black-by-SBP interactions.

**Table 2:**
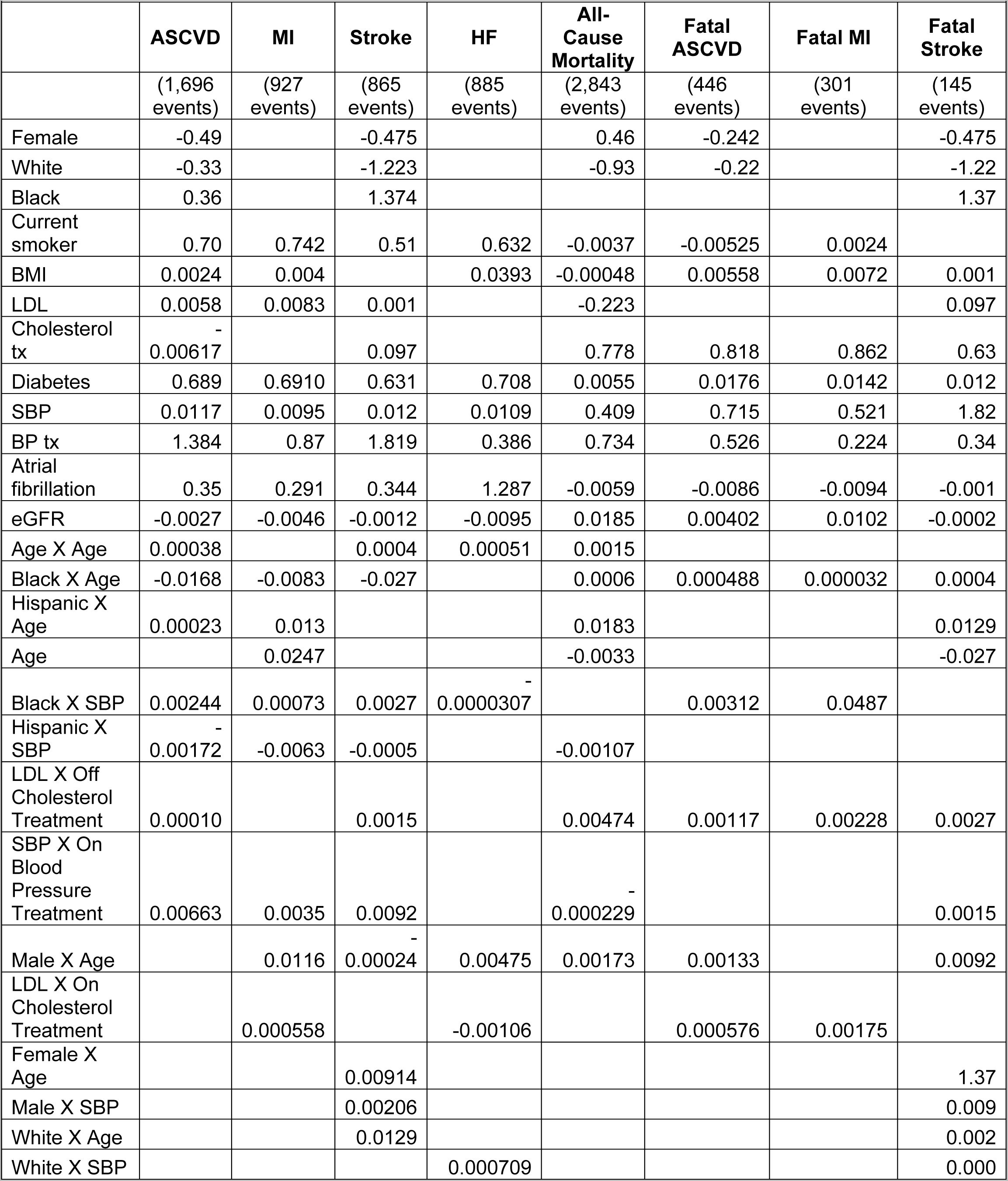
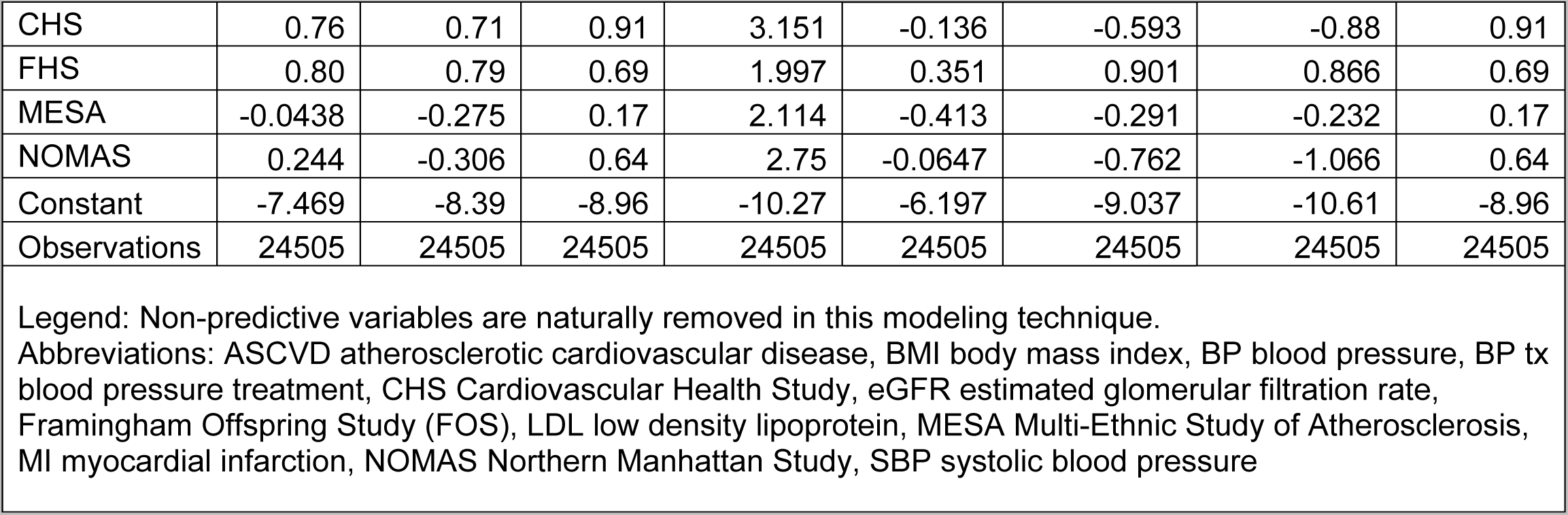
Beta-coefficients for all 10-year outcomes.

We did identify cohort-specific effects, including that participants in CHS may have had increased sensitivity to mild events as participants were more likely to have events than their risk factor profile would otherwise predict, but were less likely to die. Participants from the FHS had more fatal and nonfatal events than their risk factor profiles would otherwise predict, ARIC had fewer diagnoses of HF than would have been predicted from other studies. Table 2 contains the entire model results including the constant. By applying these results in a logistic regression formula these results could be used and replicated.

Our models’ assessments showed good predictive capacity by visual inspection of predicted probabilities (Supplemental Figures 1-8). C-statistics were between 0.745 (for MI) and 0.85 (for HF) (Table 3). Our models were well calibrated as evidenced by the excellent values for Brier Score, calibration-in-the-large, and visual assessment (Table 3 and supplementary figures 1-8). The small differences between the c-statistics of the derivation and validation shows limited overfitting, also verified by the relatively small between-run standard deviation of that difference, which was never greater than 0.04. The models show good separation, with 25^th^-75^th^ percentile results differentiating effectively for this low-risk pooled cohort. For every outcome, the bottom 25^th^ percentile threshold was below a 4% risk of an event in 10 years and the 75^th^ percentile of risk was at least three times higher. For overall ASCVD events, people at the 25^th^ percentile had a 2.7% 10-year predicted event rate and those in the 75^th^ percentile had an 8.3% rate. The observed-to-expected figures of all models are included in the supplemental appendix (Supplemental Figures 1-8). They consistently show excellent calibration for well over 90 percent of participants in all models, with substantial error occurring only in the very high-risk tails.

**Table 3:**
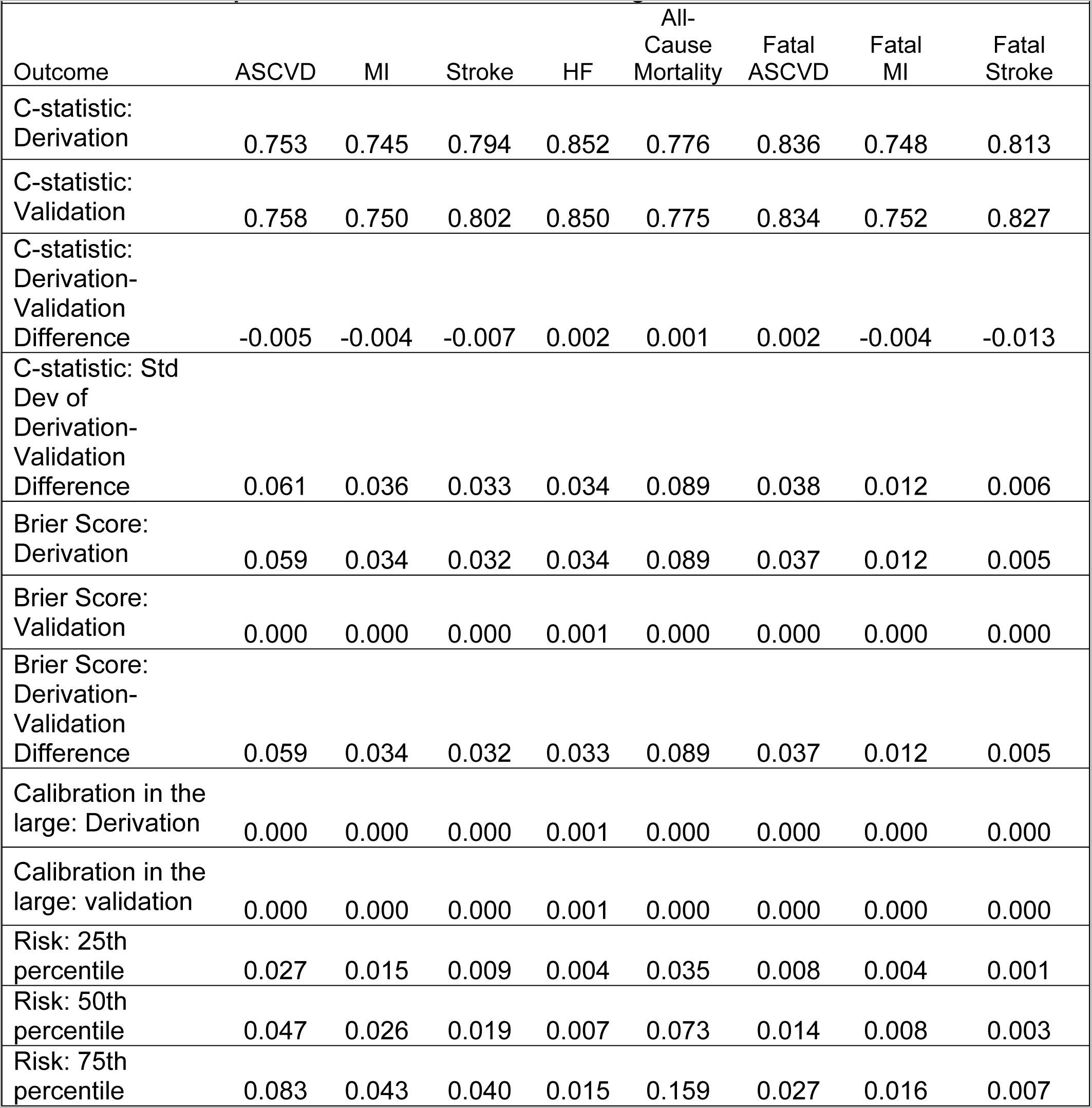
Validation statistic for every 10-year model. Each model was run 10 times on random 80% derivation samples then assessed on the remaining 20%.

| Outcome | ASCVD | MI | Stroke | HF | All-Cause Mortality | Fatal ASCVD | Fatal MI | Fatal Stroke |
| --- | --- | --- | --- | --- | --- | --- | --- | --- |
| C-statistic: Derivation | 0.753 | 0.745 | 0.794 | 0.852 | 0.776 | 0.836 | 0.748 | 0.813 |
| C-statistic: Validation | 0.758 | 0.750 | 0.802 | 0.850 | 0.775 | 0.834 | 0.752 | 0.827 |
| C-statistic: Derivation-Validation Difference | -0.005 | -0.004 | -0.007 | 0.002 | 0.001 | 0.002 | -0.004 | -0.013 |
| C-statistic: Std Dev of Derivation-Validation Difference | 0.061 | 0.036 | 0.033 | 0.034 | 0.089 | 0.038 | 0.012 | 0.006 |
| Brier Score: Derivation | 0.059 | 0.034 | 0.032 | 0.034 | 0.089 | 0.037 | 0.012 | 0.005 |
| Brier Score: Validation | 0.000 | 0.000 | 0.000 | 0.001 | 0.000 | 0.000 | 0.000 | 0.000 |
| Brier Score: Derivation-Validation Difference | 0.059 | 0.034 | 0.032 | 0.033 | 0.089 | 0.037 | 0.012 | 0.005 |
| Calibration in the large: Derivation | 0.000 | 0.000 | 0.000 | 0.001 | 0.000 | 0.000 | 0.000 | 0.000 |
| Calibration in the large: validation | 0.000 | 0.000 | 0.000 | 0.001 | 0.000 | 0.000 | 0.000 | 0.000 |
| Risk: 25th percentile | 0.027 | 0.015 | 0.009 | 0.004 | 0.035 | 0.008 | 0.004 | 0.001 |
| Risk: 50th percentile | 0.047 | 0.026 | 0.019 | 0.007 | 0.073 | 0.014 | 0.008 | 0.003 |
| Risk: 75th percentile | 0.083 | 0.043 | 0.040 | 0.015 | 0.159 | 0.027 | 0.016 | 0.007 |

Individual participants’ predicted risks for different outcomes were strongly correlated with one another, but the magnitude of correlation varied across different comparisons (Table 4). The correlation coefficient between 10-year risk of MI and stroke, as well as that between MI and all-cause mortality, was 0.68 (R^2^ = 46%). The correlation between risk of all MI and risk of fatal MI was 0.85 (R^2^ = 72%). Stroke and HF were more closely correlated with all outcomes (all-cause mortality, fatal ASCVD, and fatal stroke) than MI. The only exception is that MI was more closely associated with fatal MI than HF. The strongest correlation was between fatal MI and fatal ASCVD, with a correlation coefficient of 0.97. Fatal MI is the largest component of fatal ASCVD.

**Table 4:** Correlations between risk of 10-year outcomes.

|  | ASCVD | MI | Stroke | HF | All-Cause Mortality | Fatal ASCVD | Fatal MI | Fatal Stroke |
| --- | --- | --- | --- | --- | --- | --- | --- | --- |
| ASCVD | 1 |  |  |  |  |  |  |  |
| MI | 0.91 | 1 |  |  |  |  |  |  |
| Stroke | 0.92 | 0.68 | 1 |  |  |  |  |  |
| HF | 0.83 | 0.70 | 0.83 | 1 |  |  |  |  |
| All-Cause Mortality | 0.83 | 0.68 | 0.82 | 0.86 | 1 |  |  |  |
| Fatal ASCVD | 0.92 | 0.79 | 0.91 | 0.87 | 0.83 | 1 |  |  |
| Fatal MI | 0.92 | 0.85 | 0.85 | 0.81 | 0.79 | 0.97 | 1 |  |
| Fatal Stroke | 0.79 | 0.58 | 0.88 | 0.80 | 0.74 | 0.91 | 0.80 | 1 |

Next, we wanted to determine if participants at high risk for one outcome were meaningfully different from those at high risk for others. To do this, we identified participants who were in the top quartile for risk of MI, stroke, and all-cause mortality. In Table 5, we describe all participants who were in the top quartile for one of those three outcomes, but not the other two. Participants who were particularly high risk for MI but not stroke were disproportionately White, had higher LDL cholesterol levels and higher rates of tobacco use, and had lower rates of diabetes than those at high risk for stroke and all-cause mortality. Participants with top-quartile risk of stroke but not MI or mortality were disproportionately female and obese, had higher systolic BP levels, and were more likely to have diabetes. Participants with top-quartile risk of all-cause mortality but not of MI or stroke were strikingly older than the other high-risk groups, had lower values on all traditional ASCVD risk factors, including BP, BMI, LDL, and proportion with diabetes. Their rates of tobacco use were higher than those with high risk of stroke but lower than those with high risk of MI.

**Table 5:**
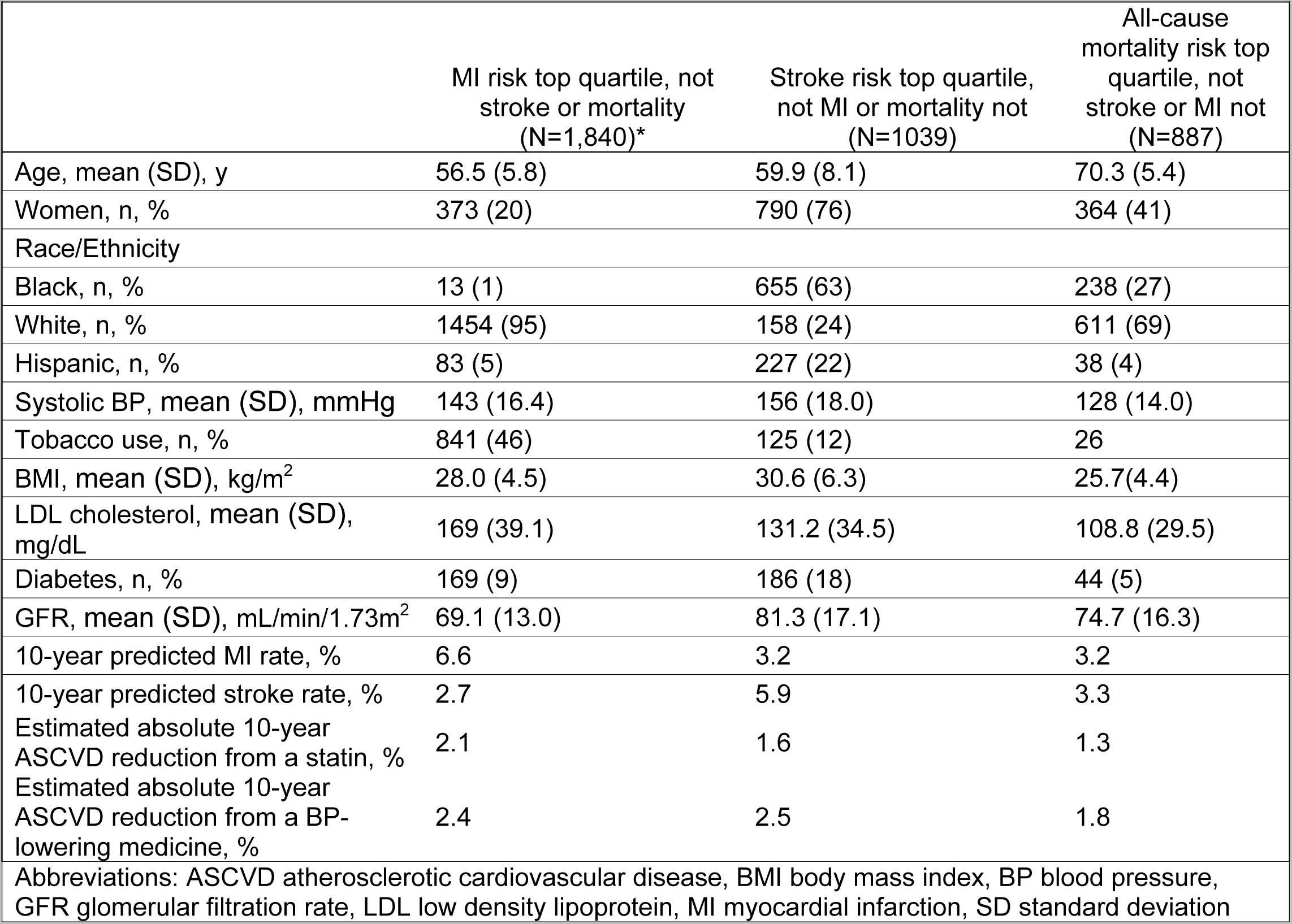
Descriptive characteristics of different high-risk groups.

| **Table 5: Descriptive characteristics of different high-risk groups** |  |  |  |
| --- | --- | --- | --- |
|  | MI risk top quartile, not stroke or mortality (N=1,840)* | Stroke risk top quartile, not MI or mortality not (N=1039) | All-cause mortality risk top quartile, not stroke or MI not (N=887) |
| Age, mean (SD), y | 56.5 (5.8) | 59.9 (8.1) | 70.3 (5.4) |
| Women, n, % | 373 (20) | 790 (76) | 364 (41) |
| Race/Ethnicity |  |  |  |
| Black, n, % | 13 (1) | 655 (63) | 238 (27) |
| White, n, % | 1454 (95) | 158 (24) | 611 (69) |
| Hispanic, n, % | 83 (5) | 227 (22) | 38 (4) |
| Systolic BP, mean (SD), mmHg | 143 (16.4) | 156 (18.0) | 128 (14.0) |
| Tobacco use, n, % | 841 (46) | 125 (12) | 26 |
| BMI, mean (SD), kg/m<sup>2</sup> | 28.0 (4.5) | 30.6 (6.3) | 25.7(4.4) |
| LDL cholesterol, mean (SD), mg/dL | 169 (39.1) | 131.2 (34.5) | 108.8 (29.5) |
| Diabetes, n, % | 169 (9) | 186 (18) | 44 (5) |
| GFR, mean (SD), mL/min/1.73m<sup>2</sup> | 69.1 (13.0) | 81.3 (17.1) | 74.7 (16.3) |
| 10-year predicted MI rate, % | 6.6 | 3.2 | 3.2 |
| 10-year predicted stroke rate, % | 2.7 | 5.9 | 3.3 |
| Estimated absolute 10-year ASCVD reduction from a statin, % | 2.1 | 1.6 | 1.3 |
| Estimated absolute 10-year ASCVD reduction from a BP-lowering medicine, % | 2.4 | 2.5 | 1.8 |
| Abbreviations: ASCVD atherosclerotic cardiovascular disease, BMI body mass index, BP blood pressure, GFR glomerular filtration rate, LDL low density lipoprotein, MI myocardial infarction, SD standard deviation |  |  |  |

We also found a difference in absolute risk reduction in events between participants who are top-quartile in risk of MI but not stroke or mortality vs. those who are top-quartile in risk of stroke but not MI or mortality when treated with a BP-lowering medications vs cholesterol lowering (Table 5). In participants who had top quartile of risk of MI, but not of stroke or all-cause mortality, one BP-lowering medication for 10 years would have an estimated 2.4% chance of preventing a first ASCVD event. A moderate-strength statin would have a 2.1% chance of preventing an event in the same period. In distinction, in participants who had top quartile risk of stroke but not MI or mortality, a BP medication would have a similar chance (2.5%) of preventing an event, but a moderate-strength statin would have a reduced chance (1.6%) chance of preventing an event. Participants with top-quartile risk of mortality but not MI or stroke had a smaller benefit for both treatments, with a 1.3% reduction in ASCVD with a moderate-strength statin and a 1.8% reduction from a BP medication.

Figure 1 also demonstrates this phenomenon. In an intermediate-risk group of 10-year ASCVD risk from 7.5% to 15%, the more a participant’s risk was due to stroke risk, the greater the benefit of BP-lowering medication. The more their risk was due to MI risk, the greater the benefit of a statin (Figure 1). Each dot represents one cohort participant.

**Figure 1:**
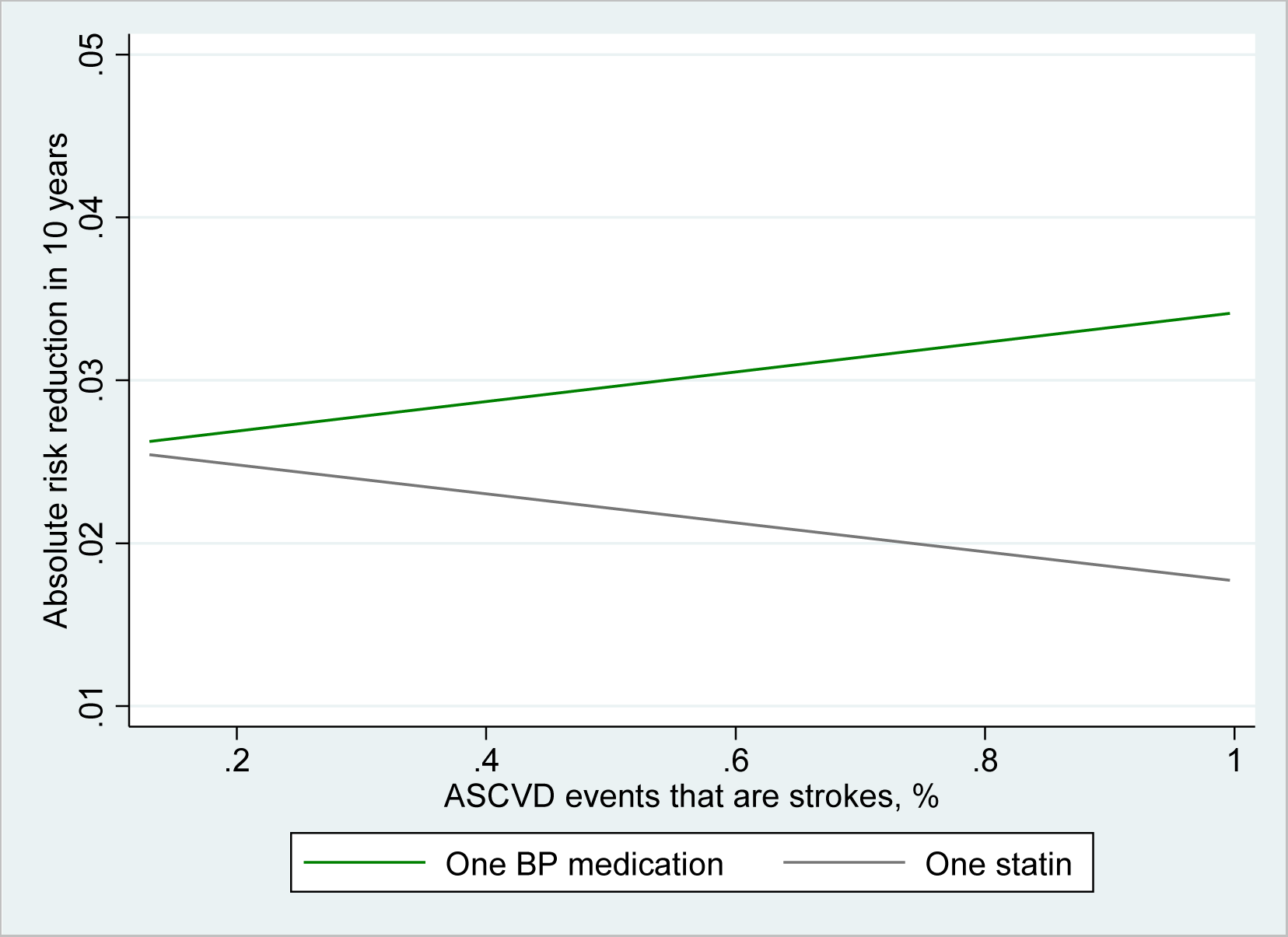
Absolute risk reduction due to one blood pressure lowering medicine vs. a moderate-strength statin in every study participant with 7.5%-15% 10-year risk by probability that their event would be a stroke. Each dot is a study participant, the lines represent the linear regression of the relationship between percentage of events that are predicted to be strokes and the absolute risk reduction of any ASCVD event in 10 years.

We performed two prespecified sensitivity analyses. One assessed the impact of using logistic regression without variable selection instead of our primary modeling technique of penalized regression and selection using ENR (Supplemental Tables S2-S5). We found that ENR had substantial benefits in quality of risk prediction, with validation c-statistics more than 0.05 better in the models predicting stroke, CHF, fatal ASCVD, and fatal stroke using ENR than logistic regression without variable selection. The correlations between the predictions were virtually unchanged, with only one comparison more than 0.05 different from the results in the primary ENR models.

The second sensitivity analysis tested the impact of using 5-year risk scores instead of 10 years (Supplemental Tables S6-S9). The effects of this were small, with validation C-statistics less than 0.02 different between 5-year and 10-year models in 6 of 8 models. We have included all models in the supplement.

## Discussion

In this study we developed risk equations for eight cardiovascular events using pooled data from five large, US cardiovascular cohort studies, first atherosclerotic cardiovascular disease (ASCVD), first fatal or nonfatal MI, first fatal or nonfatal stroke, new-onset HF, fatal ASCVD, fatal MI, fatal stroke, and all-cause mortality. We found that correlations between the risk of different types of cardiovascular disease, such as MI and stroke, were strong, usually between 0.7 and 0.88. Despite these correlations, we could identify high-risk participants who were 2.5 times as likely to have a stroke than an MI and others who were 2.5 times as likely to have an MI than a stroke. We then found that this would impact the likely benefit of taking a statin vs. a BP-lowering medication.

The risk scores we developed have advantages over previous cardiovascular risk scores. First, we developed it using multiple cohorts that had rigorous attention paid to data accuracy and case ascertainment. Our cohorts are closely related to the Pooled Cohort Equation cohorts, which allows comparison with the most common risk score used today. While risk equations exist for multiple outcomes, relatively few have allowed comparability by using the same method for each outcome.^17, 18^ We used modern techniques, including ENR, testing for overfitting with cross-validation, and assessing using modern methods such as the Brier score. Our cohorts were developed from geographically and racially diverse populations of US adults, included Hispanic adults, and included variables, such as statin treatment, that are not included in the PCEs. We evaluated the potential impact of the scores and found meaningful differences in treatment benefit among people with the same ASCVD risk.

We found that participants who were particularly high risk for one cardiovascular outcome were not necessarily high-risk in the others. Participants with high risk of MI were disproportionately White males with high LDL cholesterol levels and rates of tobacco use. Those with high risk of stroke were more likely to be Black women with high BP and BMI. This finding is consistent with studies showing greater risk of stroke in Black women than White women, a disparity that is highest at ages 50 to <60 years old, but persists after age 70.^19^ Unsurprisingly, those with a high risk of all-cause mortality were strikingly older than other high risk groups.

Finally, we found that by identifying patients who are high risk of stroke, but not MI or all-cause mortality, we could isolate patients with a greater likely benefit of BP-lowering. The greater proportion of an individual’s risk that was due to MI, the greater the benefit of statin. Participants with a risk of MI in the top quartile but not stroke or all-cause mortality had a 15% greater benefit in total ASCVD outcome from a single BP-lowering medication (2.4% vs. 2.1% 10-year reduction). But in those with a top-quartile risk of stroke but not heart disease or all-cause mortality, the risk reduction for total ASCVD was 56% greater (2.5% vs. 1.6% 10-year reduction). Participants with elevated all-cause mortality had substantially lower probability of benefitting from either drug (1.8% 10-year reduction for one BP-lowering medication and 1.3% from a statin). While not ready for clinical practice, these results shows that we could imagine personalizing care to maximize benefit based on a person’s elevated risk for a specific clinical outcome. For example, we may be more likely to consider BP reduction use in people at higher risk of stroke, due to those drug’s greater differential effectiveness in those conditions. Similarly, the glucose-lowering drugs sodium-glucose cotransporter 2 inhibitors appear to be effective at MI prevention but do not seem to reduce rates of strokes; some evidence implies the glucagon-like peptide-1 receptor -1 analogs reduce strokes more effectively than reduce MIs.^20^

In our study, each cohort had slightly different findings, particularly in overall event rates. This phenomenon, which has been seen before, demonstrates a larger concern for external validity in all predictive model research.^15, 21^ We addressed this by removing the cohort effect and normalizing the Y-intercept. This minimizes multiple biases but there is no way to be certain that between-cohort differences could have created effects seen only in specific variables. The data in this study were obtained at different times in many locations across the United States. From the time of data acquisition, many care practices have changed, most dramatically an increased rate of statin use and continued decline in tobacco use. Almost half of our data is from the ARIC cohort.

Another limitation is the non-causal nature of risk scores and the subjectivity in developing them. Higher risk participants will not necessarily receive more benefit from treatment, though existing research indicates they usually do.^23–25^ Our results would have changed slightly with different analytic choices, including the unavoidably subjective nature of which potential predictor variables to include. We opted to use variables that have been used many times in cardiovascular prediction and are easy to obtain clinically. Some potential predictors, such as use of newer diabetes medicines, were not included because data from the cohorts was not recent enough.

Our work shows that it is valuable to predict cardiovascular outcomes independently, while also providing the tools to do so. These results have many possible utilities. They could be used clinically by participants who are particularly concerned about one event type over another. They could be used in cost effectiveness studies and policy simulations to guide the accuracy of using treatments that are more effective at preventing one event type vs. another. They could also help guide population health interventions. Future work should include understanding how much integrating these findings into clinical and public health practice can influence outcomes.

## Data Availability

Data referred to in this manuscript is owned by each respective study cohort. Data will be made available by request with proper data sharing agreements in place with the cohorts' institutions.

## Acknowledgements

This research project is supported by a grant R01 NS102715 from the National Institute of Neurological Disorders and Stroke (NINDS), National Institutes of Health, Department of Health and Human Service. The NINDS was not involved in the design and conduct of the study; collection, management, analysis, and interpretation of the data; preparation, review, or approval of the manuscript; and decision to submit the manuscript for publication except one representative (author RFG) of the funding agency reviewed the manuscript. The content is solely the responsibility of the authors and does not necessarily represent the official views of the National Institute of Neurological Disorders and Stroke or the National Institutes of Health.

This research was supported by contracts HHSN268201200036C, HHSN268200800007C, HHSN268201800001C, N01HC55222, N01HC85079, N01HC85080, N01HC85081, N01HC85082, N01HC85083, N01HC85086, 75N92021D00006, and grants U01HL080295 and U01HL130114 from the National Heart, Lung, and Blood Institute (NHLBI), with additional contribution from the National Institute of Neurological Disorders and Stroke (NINDS). Additional support was provided by R01AG023629 from the National Institute on Aging (NIA). A full list of principal CHS investigators and institutions can be found at **<u>CHS-NHLBI.org</u>**. The content of this manuscript is solely the responsibility of the authors and does not necessarily represent the official views of the National Institutes of Health

The Framingham Heart Study (FHS) is conducted and supported by the National Heart, Lung, and Blood Institute (NHLBI) in collaboration with Boston University (Contract No. N01-HC-25195, HHSN268201500001I and 75N92019D00031). This manuscript was not prepared in collaboration with investigators of the Framingham Heart Study and does not necessarily reflect the opinions or views of the Framingham Heart Study, Boston University, or NHLBI.

The Multi-Ethnic Study of Atherosclerosis study (MESA) is conducted and supported by the National Heart, Lung, and Blood Institute (NHLBI) in collaboration with MESA investigators. Support for MESA is provided by contracts N01-HC95159, N01-HC-95160, N01-HC-95161, N01-HC-95162, N01-HC-95163, N01-HC-95164, N01-HC-95165, N01-HC95166, N01-HC-95167, N01-HC-95168, N01-HC-95169 and CTSA UL1-RR-024156.

The Atherosclerosis Risk in Communities study has been funded in whole or in part with Federal funds from the National Heart, Lung, and Blood Institute, National Institutes of Health, Department of Health and Human Services, under Contract nos. (75N92022D00001, 75N92022D00002, 75N92022D00003, 75N92022D00004, 75N92022D00005). Additionally, Dr. Gottesman was supported by the NINDS intramural research program. The authors thank the staff and participants of the ARIC study for their important contributions.

The Northern Manhattan Stroke (NOMAS) study has been funded at least in part with federal funds from the National Institutes of Health, National Institute of Neurological Disorders and Stroke by R01 NS29993, AG066162 and AG057709.

